# Impact of the COVID-19 pandemic on the mental health of Greek adults: a cross-sectional survey

**DOI:** 10.1101/2021.02.20.21252129

**Authors:** Efthimios Dragotis, Korina Atsopardi, Anastasia Barbouni, Konstantinos Poulas, Konstantinos Farsalinos

## Abstract

**Objectives:** The coronavirus disease 2019 (COVID-19) pandemic and mitigation measures based on social distancing are expected to have serious adverse effects on mental health. This cross-sectional study aimed to examine self-reported changes in the mental health status of Greek adults.

**Study design:** The current study is a primary research conducted on Greek adults during the first wave of the epidemic (March to April 2020).

**Methods:** A total of 527 individuals participated in an online survey using a validated questionnaire (State-Trait Anxiety Inventory-STAI and DASS-21).

**Results:** The respondents had a moderate mental health status based on the following scores: STAI-S, 45.8; STAI-T, 40.7; depression, 4.6; anxiety, 3.1; and stress, 6.1. Women, younger respondents, those from lower income households, and those living in smaller apartments experienced increased depression, anxiety, and stress. Additionally, infection control practices during the COVID-19 pandemic such as the use of masks, gloves, and antiseptic can drastically decrease the prevalence of mental health illnesses.

**Conclusions:** These findings can be used by the Greek State to reduce the effects of COVID-19 on the mental health of the population and protect socially vulnerable groups.

## Introduction

The coronavirus disease (COVID-19) was first reported in Wuhan (Hubei Province, China) in late December 2019, and within a few months, it was declared a pandemic by the World Health Organization on March 11, 2020. On February 26, 2020, the first COVID-19 case was reported in Greece. The government immediately implemented social distancing measures and, on March 23, 2020, a nationwide horizontal lockdown was imposed. Such restrictive measures disrupt social and professional life and are known to have negative psychological effects, such as confusion, anger, and post-traumatic distress [1,2]. Anxiety, stress, depression, and post-traumatic stress have also been observed in previous viral disease outbreaks, such as severe acute respiratory syndrome, middle East respiratory syndrome, Ebola, and H1N1 outbreaks [3–11].

Factors such as fear of a novel disease with limited understanding, as well as media reports about COVID-19 patients and deaths, create an environment of increased anxiety and fear [12,13], which COVID-19 patients may also experience [14,15]. Moreover, social distancing measures may result in feelings of disappointment, irritability, loneliness, denial, and depression [1,15]. Studies in China and Turkey reported that lockdown measures increase the levels of anxiety and depression in the general population [16–18]. Health professionals are also experiencing intense physical and psychological pressures [19,20]. Studies in specific population subgroups in Greece reported that children, high school students, university students, and patients with chronic disease experienced elevated stress during the national lockdown [2,21– 24]. This study aimed to examine the impact of the pandemic and horizontal social distancing measures implemented in Greece on the mental health of Greek adults.

## Methods

This cross-sectional survey was performed from May 2 to 5, 2020, during the last few days of implementing stay-at-home measures in Greece since March 23. A questionnaire was prepared and validated in 15 adults (who did not participate in the study) using the cognitive interviewing method [25]. This was divided into six sections. The first section included demographics such as sex, age group, type of place of residence, education level, citizenship, work status, income level, and family status. The second section included questions about symptoms compatible with COVID-19 that were experienced by the participants during the period from February to May 2020. The purpose of these questions was to examine whether individuals with such symptoms had experienced different mental health effects from those without symptoms. The following symptoms were recorded: fever ≥37 °C, cough, headache, nasal congestion, sore throat, shortness of breath, malaise, fatigue, olfactory and gustatory disturbances, and gastrointestinal symptoms. The third section included questions that examined the mental health status of the respondents. Stress in the working environment was measured using the State-Trait Anxiety Inventory (STAI) questionnaire [26], which consists of 40 statements to which participants are asked to respond using a 4-point Likert scale. This questionnaire examines two types of anxiety: the first is state anxiety, anxiety about an event, and trait anxiety; and the second is the anxiety level, which are considered personal characteristics. For the first 20 questions (transient stress), the Likert scale ranges from “Not at all” to “Too much,” while for the next 20 questions (permanent stress), the scale ranges from “Almost never” to “Almost always.” The Depression Anxiety and Stress Scale (DASS-21) was used [27], which is a short version (21 items) of a 42-item self-report instrument designed to measure three related negative emotional states: depression, anxiety, and tension/stress [27].Again, a 4-point Likert scale was used, ranging from 0 – “did not apply to me at all” to 3-“Applied to me very much, or most of the time” These questionnaires have been used in other studies evaluating mental health status during the COVID-19 pandemic [28–33].

## Results

### Sample demographics

The majority of the sample (98.7%) comprised Greek respondents. The sample consisted of 64% female and 36% male respondents. The prevalent age group was 35–44 years (24.5%) and 25–34 years (23.8%). Majority of the respondents were living in cities (80%) and in medium-sized apartments from 50 to 100 m^2^ (52.5%). Nearly half of the respondents (47.5%) were graduates. They worked in different sectors: 16.8% were public sector employees, 28.8% were private sector employees, 19% were freelancers, 7.6% were retired, 11.8% were unemployed, and 16.0% were students. Majority of them (86.6%) did not work as health professionals. Approximately 34.1% were not working from home (type A of document). The respondents had different income levels: 28.2 % had no income, while 28.7% had over 1000 euros. Of the respondents, 42% were married, and 49% were unmarried. Majority (57%) had no dependent family members. Lastly, the 18% was living in an apartment up to 50 m^2^, the 52,5% in an apartment up to 100 m^2^ and the 29,5% in apartments over 100 m^2^.

### Anxiety, stress, and depression

Measuring depression, anxiety, and stress levels. Table 2 shows that the respondents had a moderate level of mental health status.

**Table 1.**
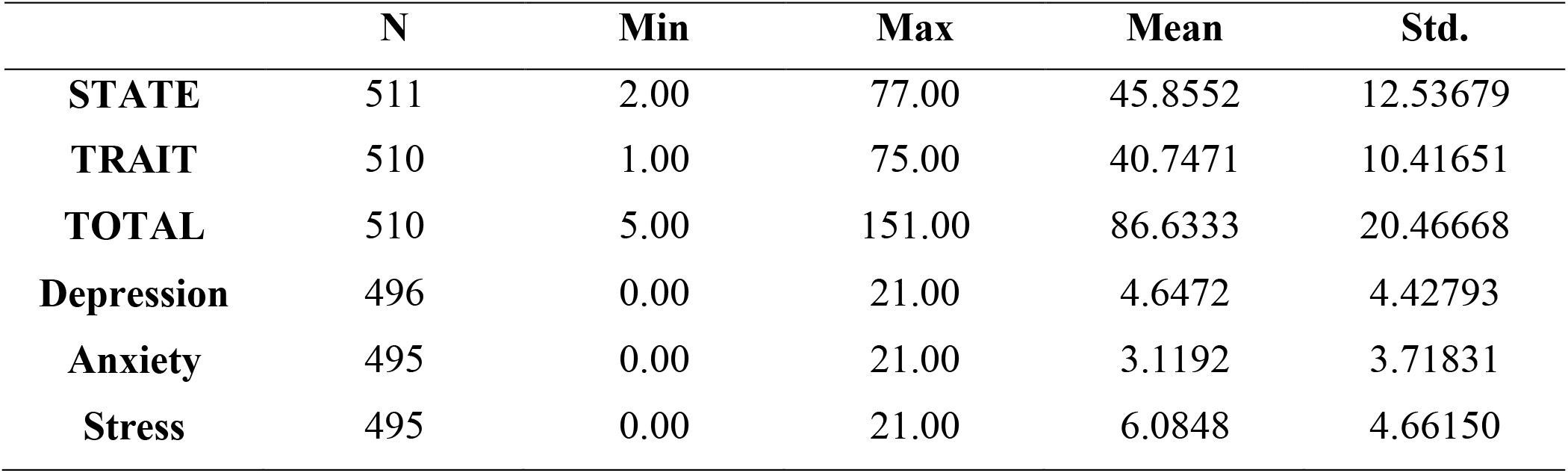
Anxiety, stress, and depression

|  | **N** | **Min** | **Max** | **Mean** | **Std.** |
| --- | --- | --- | --- | --- | --- |
| **STATE** | 511 | 2.00 | 77.00 | 45.8552 | 12.53679 |
| **TRAIT** | 510 | 1.00 | 75.00 | 40.7471 | 10.41651 |
| **TOTAL** | 510 | 5.00 | 151.00 | 86.6333 | 20.46668 |
| **Depression** | 496 | 0.00 | 21.00 | 4.6472 | 4.42793 |
| **Anxiety** | 495 | 0.00 | 21.00 | 3.1192 | 3.71831 |
| **Stress** | 495 | 0.00 | 21.00 | 6.0848 | 4.66150 |

**Table 2.**
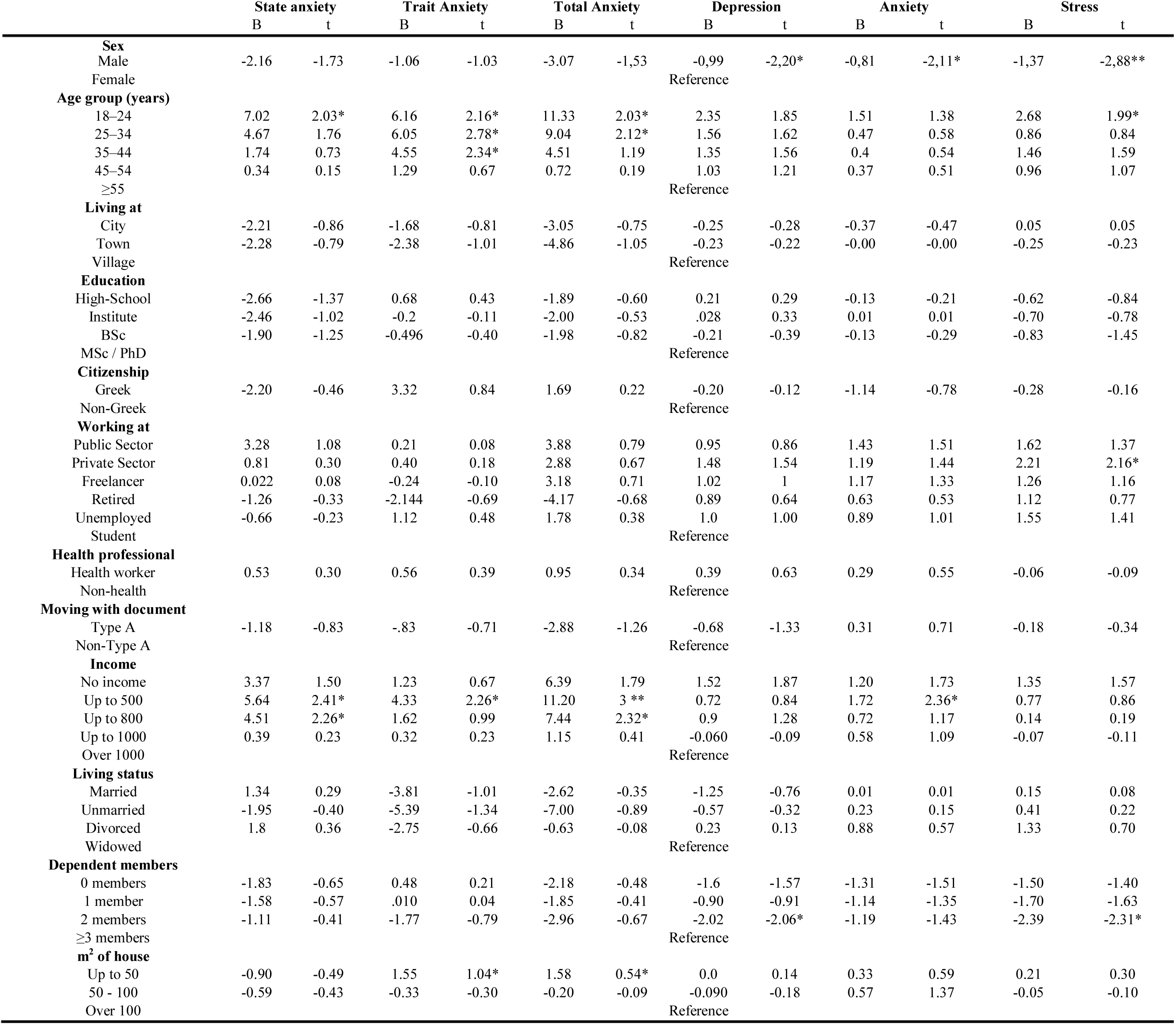
Association between demographics and mental health status

### Depression, anxiety, and stress not related to COVID-19

The respondents’ demographics and medical histories were analyzed to determine their relationship with mental health. The results (Table 3) indicate that women presented with significantly higher mean scores for stress, depression, and anxiety; while the 18–24 year old group (the younger sample group) presented with significantly higher mean scores for anxiety and stress, with the group that worked in the private sector presenting with higher stress levels. Moreover, people with two dependent family members had significantly higher trait stress and depression than the other groups in the sample. Furthermore, the subgroups with income up to 500 euros and living in an apartment with an area of up to 50 m^2^ presented with significantly higher anxiety symptoms. As for medical history, results indicate that mental health issues were significantly higher in only four cases: people with asthma, who had higher state anxiety; people with cancer, who had higher trait anxiety; people with immunosuppression, who had higher anxiety; and people with hypertension, who had stress.

**Table 3.**
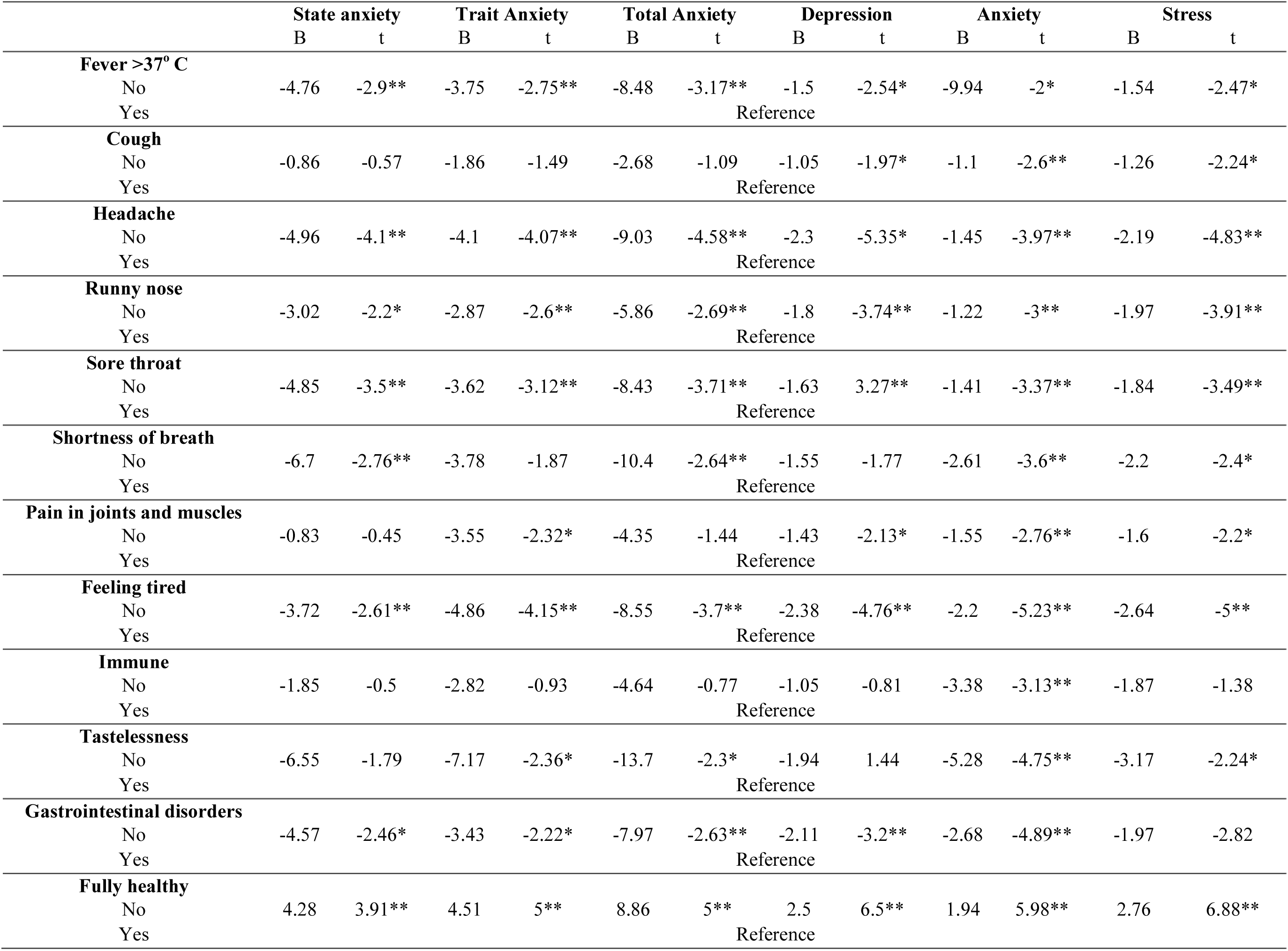
Association between symptoms related to COVID-19 andmental health status

### Depression, anxiety, and stress related to COVID-19

This study evaluated the effect of different infection control practices taken during the spread of COVID-19 on stress, depression, and anxiety levels. There was a statistically significant decrease in anxiety levels with the use of masks (trait anxiety, total anxiety, and anxiety), antiseptic (state anxiety and anxiety), and gloves (anxiety). People who used antiseptics, wearing masks, and gloves more often tended to have lower levels of anxiety. Moreover, significantly higher depression levels were seen in people who share their utensils more often. The following table summarizes the correlations that were observed to be statistically significant. The statements “cover mouth on sneeze,” “hand washing,” “washing hands after cough,” “washing hands after touching contaminated objects,” and “products washing” do not have statistically significant correlations with mental health illnesses.

Furthermore, to evaluate the effect of COVID-19 potential symptoms on mental health levels, 12 symptoms/ conditions were tested. The results (Table 4) indicate that those who presented with each symptom had significantly higher levels of stress, depression, and anxiety than those who did not. Specifically, people who had fever at 37 °C and above, cough, headache, runny nose, sore throat, pain in their joints and muscles, and felt tired had significantly higher mean scores for depression, stress, and anxiety. In addition, the sample with gastrointestinal disorders and immunity presented with higher levels of anxiety. Finally, the sample with shortness of breath and tastelessness presented with significantly higher mean scores for depression. At the same time, the sample that declared themselves healthy presented with significantly lower mean scores for depression, stress, and anxiety.

**Table 4.** Levels of mental health illnesses

| **Level / Mental health illness** | **Depression** | **Anxiety** | **Stress** |
| --- | --- | --- | --- |
| Normal | 60% | 66.5% | 64.8% |
| Mild | 11.5% | 13.1% | 12.4% |
| Moderate | 12.5% | 6.7% | 11.1% |
| Severe | 10% | 6.7% | 9.3% |
| Extremely Severe | 5% | 6.7% | 2.4% |

## Discussion

### Main findings of the study

The results show that the study population had a sufficient level of knowledge about the virus in terms of its transmission and their vulnerability to it. People with health problems as well as the elderly people are at increased danger in the case of coronavirus infection. Recent studies and clinical data have shown that mortality rates are increased for people 55 and above, as well as for people with an underlying disease. The respondents seemed to be well-informed about the modes of virus transmission, and their most common practices included covering the mouth when coughing or sneezing, washing their hands with soap and water, and using a face mask in public places. Majority of the respondents (54.3%) reported that from February to May, one or more symptoms related to COVID-19 were present.

### What is already known at this topic

Considering work anxiety levels (STAI), the sample reports a STAI-S score of 45.85 and a STAI-T score of 40.74. Relevant research has evaluated either higher [31] or lower [34] STAI-S and STAI-T scores. Our study revealed medium anxiety levels (STAI-S: 32.7% low, 54.2% moderate, 12.3% severe – STAI-T: 47.2% low, 47.2% moderate, and 4.1% severe), which are similar to those of other studies. Moderate to severe levels were seen in 66.5% (STAI-S) and 52% (STAI-T) of the respondents, respectively, which is a high percentage when compared to results from relevant literature [35]. As for the rest of the mental health illnesses (DASS-21), the levels of depression, anxiety, and stress were 4.65, 3.12, and 6.08, respectively. These results represent lower levels of mental health illnesses compared to those of previous studies conducted in different places within the same research period. Odriozola-Gonzáleza et al. [36] reported levels of depression, anxiety, and stress to be 5.52, 3.34, and 6.81, respectively in Spain. According to Wang et al. [30] there are significant differences between DASS-21 scores in Europe (Poland) and Asia (China), but these two countries still have higher scores than Greece (China - Depression: 6.25, Anxiety: 6.16 and Stress: 7.76; Poland - Depression: 10.06, Anxiety: 7.65 and Stress: 14.00). According to Verma et al. [37] the levels of depression, anxiety, and stress is 8.39, 6.53, and 8.83, respectively in India. Although the above findings signify a higher level of mental health illness than our results, our sample had a relatively high score on the severe and extremely severe DAS scales. The scores of the sample were classified into five categories: normal, mild, moderate, severe, and extremely severe. Majority of the current study’s respondents (60%–65%) demonstrated normal DASS-21 levels; however, these scores are relatively lower than those in the current literature. Mild and moderate levels were seen in approximately 20% and 24% of the sample, respectively, which is higher than that those seen in other studies. Finally, severe and extremely severe levels were seen in approximately 11.2% and 15% of the sample, respectively, which is significantly higher than those seen in other studies [28,29,33,38–40].

This study aimed to assess potential significant differences between subgroups of the sample in order to evaluate the factors that have a significant role in mental health status, which were found to be sex, age, income level, and place of living size. The female subgroup had significantly higher depression, stress, and anxiety levels; younger respondents had significantly higher state and trait anxiety levels; low-income workers had significantly lower anxiety, state, and trait anxiety levels; and the individuals that lived in a place with an area less than 50 m^2^ had significantly higher trait anxiety scores. These findings are similar to other studies conducted within the period of the COVID-19 outbreak in Europe, with female subgroups reported to have significantly higher anxiety, stress, and depression levels [18,39–47]. Another group with increased depression, anxiety, or stress levels, as reported in related literature, is the younger subgroup [28,29,31,34,41,42,47] Our findings confirm this significant difference. A third factor that is strongly linked with higher levels of depression, anxiety, or stress is income. Our findings support conclusions from related literature that low-income and unemployed people have higher levels of DASS during the current period [31,34,35,40,44,47]. Some researchers also point out the significantly decreased depression, anxiety, and stress levels in single/unmarried groups [33,39,44] however, our research did not evaluate significant differences within different family status subgroups. Furthermore, our study did not reveal significant differences in depression, anxiety, or stress (normal, state, or trait) levels between healthcare professionals and other community samples, in contrast to relevant literature findings stating that healthcare workers have higher depression anxiety and stress levels [34,40,44].

The results of our research show that the link between specific measures of protection and mental health illnesses is strong. The use of masks, antiseptics, and gloves was found to be vital to the mental health of the respondents. The use of each of these decreases the levels of specific mental health illnesses. These results are supported by current literature findings. Within the relevant literature, use of a mask and gloves as a means of protection are reported to generate significantly lower scores for mental health illnesses [28,29,39,47]. Moreover, people who wash their hands more often, specifically after coughing or sneezing, report lower levels of depression, anxiety, or stress levels [33,39,40,47].

Another significant finding is that the specific symptoms of COVID-19 can create serious mental health issues, as people with the abovementioned symptoms have a significantly higher level of depression, anxiety, stress, state, and trait anxiety. The results of our research support the relevant literature findings that disclose a significant relation between mental health illnesses and symptoms of COVID-19: the people who had one or more symptoms related to COVID-19 had significant higher levels of Depression, Anxiety and Stress [28–30,33,39,40,47]. During the outbreak, people with any symptoms that could be related to COVID-19 were suspected to be positive for the virus. The suspicion of being ill with a severe virus such as COVID-19 is considered a risk factor for depression, anxiety, and stress.

### What the study adds

The serious effect of COVID-19 and restrictive social distancing measures on the mental health of specific groups is an important finding of this study. Relevant literature findings and the current study results show adverse mental health impacts of the COVID-19 outbreak and the period of the lockdown. However, the impact on specific subgroups, namely women, individuals living in a smaller apartment (under 50 m2), individuals with low-income, and the younger population, is significantly higher; they were found to experience increased levels of depression, anxiety, and stress. Moreover, the results of the DASS-21 showed higher mean scores and percentages of “severe” and “extremely severe” mental health illnesses, which means that people with these mental health issues experienced them more intensely. Lastly, the results suggest that there are factors that have a moderating role in the relationship between the COVID-19 outbreak, restrictive measures, and mental health status. Such factors include the use of a mask, gloves, and antiseptics, which significantly decrease mental health illnesses such as anxiety and stress. The results of our study show that, except for their commonly accepted medical significance (protecting from the spread), these infection control practices also have mental health significance. Based on the abovementioned results and the relevant findings of the current literature, the authorities must focus on the most vulnerable groups and take specific measures for their medical and mental health status. The COVID-19 outbreak demands the enactment of targeted care and strengthening public health by focusing on every subgroup, specifically the most vulnerable, in contrast to merely taking horizontal measures that generally affect the entire population. Restrictive measures, specifically strict, prolonged, and recurrent ones, might lead to serious mental health issues.

### Limitations of the study

The study has several strengths that are related with the contribution to the relevant knowledge and the correlation of the findings with the prior literature. However, some main limitations have to be referred as factors that may question the objectivity of the research. The current research is carried out through online survey promoted through social media. This is a non-probability methodological approach as specific groups (e.g., age groups, economic status groups, etc.) of the population have more chances to become part of the research sample. Another limitation is concerning a demographic variable and its meaning. The apartment of living capacity can be an economic or social status indicator; however, it is not the unique one. Even if the findings are supported by the low-income question, the apartment capacity is not an economic indicator by itself.

## Data Availability

All data are available

